# Network meta-analysis on the effects of various exercise modalities on pain control in populations with chronic nonspecific low back pain

**DOI:** 10.1101/2024.11.07.24316901

**Authors:** Ke Zhao, Hua Li, Li Li, Yongxiao li

## Abstract

**Objective:** This study aims to comprehensively evaluate and compare the effects of various exercise modalities on the control of chronic nonspecific low back pain through a network meta-analysis, in order to identify the most effective interventions.

**Methods:** Adhering strictly to the PRISMA guidelines, this study selected randomized controlled trials from databases including PubMed, Web of Science, Embase, Cochrane Library, and Scopus, up to June 30, 2024. Data were processed using Stata 17.0 software, and the effect sizes were synthesized using standardized mean differences (SMD) with 95% confidence intervals (CI). The SUCRA method was used to rank the effectiveness of the interventions.

**Results:** A total of 26 studies involving 1,507 participants aged between 20 and 63.5 years were included. The network meta-analysis revealed that yoga [SMD = −1.71 (−2.93, −0.49), P < 0.05] and core stability training [SMD = −0.81 (−1.44, −0.18), P < 0.05] were significantly more effective than the control group. SUCRA probability ranking indicated that Tai Chi (SUCRA = 77.4) might be the best modality for improving control of chronic nonspecific low back pain.

**Conclusion:** This network meta-analysis demonstrates the intervention effects of different exercise modalities on chronic nonspecific low back pain, with Tai Chi potentially being the most effective intervention. This provides an important reference for non-pharmacological interventions in chronic nonspecific low back pain.

---

Chronic nonspecific low back pain, as the main type of back pain, is characterized by unclear pain causes and the absence of specific pathological structural changes. It primarily manifests as soreness in the lower back or lumbosacral area, which may be accompanied by limited mobility[1, 2]. In 2020, 619 million people globally were affected by back pain, a number expected to rise to 843 million by 2050[3]. In the United States, chronic nonspecific low back pain has become one of the most common health issues among adults, with millions affected annually, resulting in substantial medical expenses, as reported by the Centers for Disease Control and Prevention (CDC)[4]. Chronic nonspecific low back pain not only causes persistent pain but also adds to the medical burden, posing a severe public health issue[5, 6]Therefore, enhancing the timely diagnosis, comprehensive treatment, and prevention of chronic nonspecific low back pain is of great importance and has become a significant challenge in the global public health field [7].

While pharmaceutical treatment, a traditional method, may alleviate pain in the short term, its effects are often limited and come with a range of side effects, such as dependency, gastrointestinal discomfort, and cardiovascular risks [8].Hence, seeking alternative therapies to alleviate patient suffering and improve quality of life is crucial. Against this backdrop, exercise therapy has attracted widespread attention as a potentially more effective intervention. According to a review by Hayden, exercise therapy may be more effective than conventional treatments in improving pain [9]. As research in exercise science deepens, physical exercise, as a safe and economical intervention, has shown significant effects on various types of chronic pain[10–12]. Beyond alleviating pain, exercise also effectively improves patients’ mental health[13]. For example, a randomized controlled trial indicated that chronic nonspecific low back pain patients who regularly engage in aerobic exercise outperformed the control group in terms of pain relief, providing strong evidence for the efficacy of exercise as an intervention[14]. There are many types of exercise measures used to intervene in chronic nonspecific low back pain, such as yoga, combined exercises, Pilates, Qigong therapy, suspension training, and Tai Chi, all of which have shown positive therapeutic effects[15–17].

However, there is still considerable debate over which type of exercise intervention is most effective. Traditional meta-analyses can only merge studies that compare directly, making it difficult to analyze data without direct comparisons[17–19]. Network meta-analysis, on the other hand, can merge both direct and indirect evidence to compare different interventions, thus addressing the limitations of traditional meta-analyses in indirect comparisons and offering greater precision [20].Therefore, this article, based on published studies, compares the efficacy of different exercise modalities on chronic nonspecific low back pain through network meta-analysis, providing a comprehensive and reliable reference for selecting the best intervention for chronic nonspecific low back pain through direct comparison.

## 1 Data and Methods

This systematic review was conducted in accordance with the Preferred Reporting Items for Systematic Reviews and Meta-Analyses: PRISMA Statemen[21]. and registered on the Prospective Register of Systematic Reviews (PROSPERO) platform with the registration number CRD42024597460.

### 1.1 Search Methods

Searches were conducted in five databases: EBSCO, PubMed, Web of Science, Embase, and Cochrane, with the search period extending up to June 30, 2024, starting from the inception of each database. The search strategy involved a combination of the following keywords: (1) exercise, Strength Training, physical exercise, physical activity, Pilates, sports, fitness, Functional Training, Cardio Training, Yoga, Exercise Therapy; (2) Adult, Mature, Grown-up, Adulthood, Middle-aged, Elderly, Senior, Full-grown, Professional, Independent, Mature Individual, Established Person; (3) Chronic Non-Specific Low Back Pain, CNSLBP, Nonspecific Low Back Pain, Chronic Low Back Pain, Persistent Low Back Pain, RCT, experiment, trial. The Boolean operator “AND” was used to link these three groups of terms. Additionally, references from published systematic reviews and meta-analyses were tracked and included in the study to ensure the comprehensiveness of the literature search. For example, the search string used in Web of Science was: TS=(exercise OR “Strength Training” OR “physical exercise” OR “physical activity” OR Pilates OR sports OR fitness OR “Functional Training” OR “Cardio Training” OR Yoga OR “Exercise Therapy”) AND TS=(Adult OR Mature OR “Grown-up” OR Adulthood OR “Middle-aged” OR Elderly OR Senior OR “Full-grown” OR Professional OR Independent OR “Mature Individual” OR “Established Person”) AND TI=(“Chronic Non-Specific Low Back Pain” OR CNSLBP OR “Nonspecific Low Back Pain” OR “Chronic Low Back Pain” OR “Persistent Low Back Pain”).

### 1.2 Inclusion Criteria

Based on the PICOS principles, criteria for inclusion and exclusion of literature were established. The inclusion criteria are as follows: (1) The study population consists of adults (aged 18 and above) suffering from nonspecific low back pain; (2) Interventions involve different modes of exercise in a real-world setting, with an intervention period of no less than 4 weeks; (3) Interventions include conventional treatment methods; (4) Treatment outcomes are recorded using the Visual Analogue Scale (VAS); (5) The study design is a randomized controlled trial (RCT). The exclusion criteria are as follows: (1) Cross-sectional studies, case-control studies, and other descriptive studies; (2) Reviews, abstracts, letters, and comments lacking a clear description of the study design; (3) Articles with incomplete data provision and where required data cannot be obtained through other means.

### 1.3 Data Extraction

Data extraction and literature screening were independently conducted by two members of the research team, both trained in evidence-based methodology and with long-term focus on chronic nonspecific low back pain in adults. In cases of disagreement during the screening or extraction process, a third member of the team, experienced in the field of treatment for chronic nonspecific low back pain in adults, was consulted. Together, they discussed and made the final decisions. The process primarily focused on extracting key information including the first author’s name, year of publication, country where the study was conducted, sample size of the adult population involved in the study, type of interventions used, duration of the intervention, frequency of the intervention, complete intervention cycle, and the primary outcome measures used to assess the effectiveness of the intervention.

### 1.4 Research Quality Assessment

The quality of the studies included in the analysis was comprehensively and rigorously assessed using the Cochrane Risk of Bias Assessment Tool[22]. This assessment framework thoroughly covers seven core dimensions: effectiveness of the randomization process, blinding of participants and personnel, blinding of outcome assessors, concealment of allocation, completeness and accuracy of outcome data, selective reporting of study results, and the presence of other potential biases. For each dimension, a detailed evaluation was conducted and the results were categorized into three levels: high risk of bias, indicating a significant potential for bias; unclear risk of bias, where risk levels cannot be determined due to insufficient information; and low risk of bias, suggesting lower risk and higher reliability of data.

### 1.5 Data Processing

Data processing was conducted using Stata 17.0 software. As the outcome measures of the studies were continuous variables and the assessment tools and units varied slightly among individual studies, the standardized mean difference (SMD) combined with a 95% confidence interval (CI) was used to calculate the combined effect size. The comparison results were interpreted based on effect size (ES) and the 95% confidence interval (95% CI), and interventions were ranked using both the magnitude of ES and the Surface Under the Cumulative Ranking curve (SUCRA) values. Additionally, a corrected comparison funnel plot was utilized to assess publication bias.

## 2. Research Results

### 2.1 Literature Search Results

A preliminary search from various databases yielded 1070 relevant publications. Using a literature management software and by reviewing titles and abstracts, publications that did not meet the inclusion criteria were excluded. Ultimately, 26 studies were included in the analysis, with the specific selection process illustrated in Figure 1.

### 2.2 Basic Characteristics of the Included Studies

This research included 26 articles, encompassing 1,507 participants with age ranging from 20 to 63.5 years. The basic characteristics of the studies included are presented in Table 1.

**Table 1.**
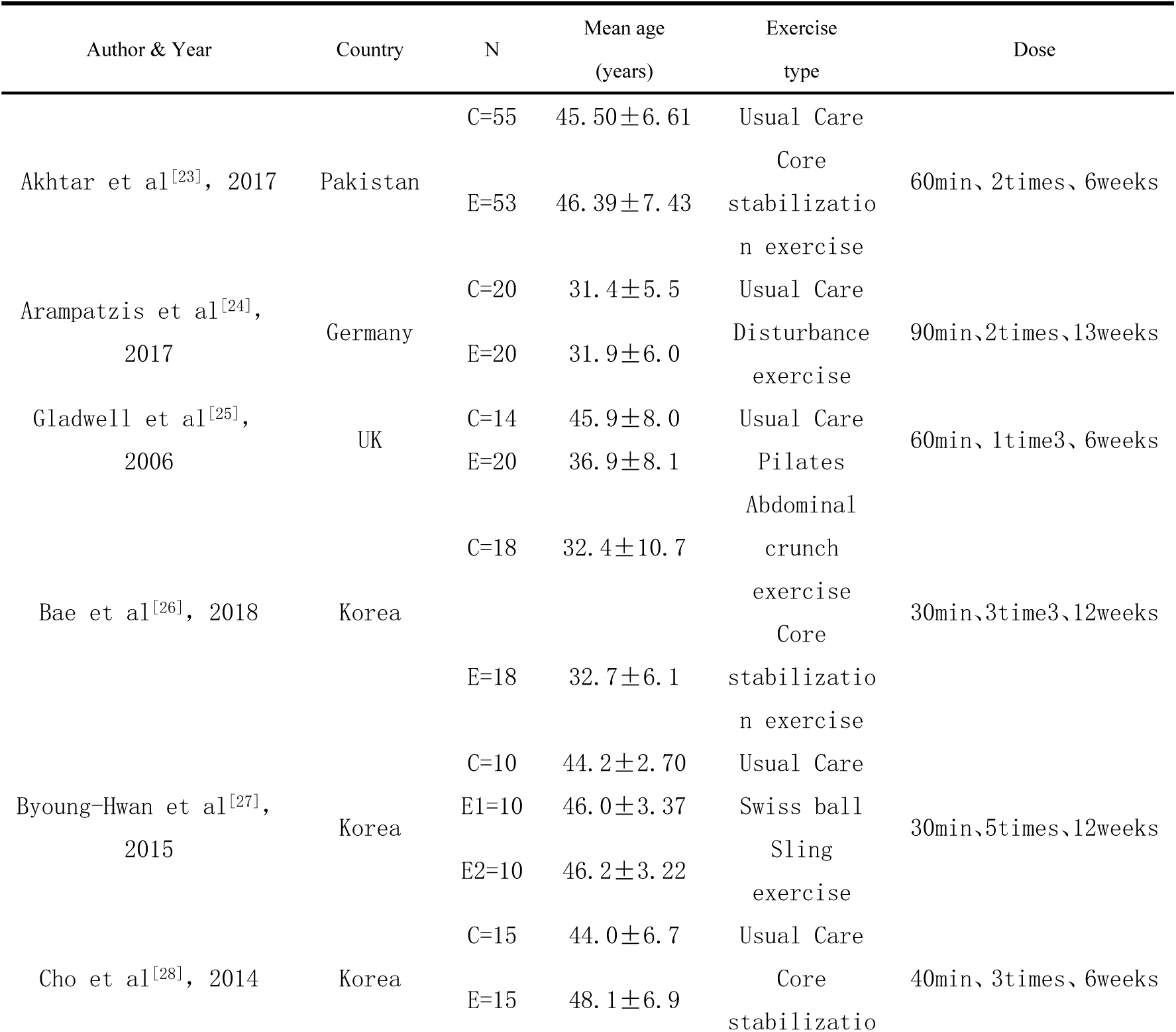
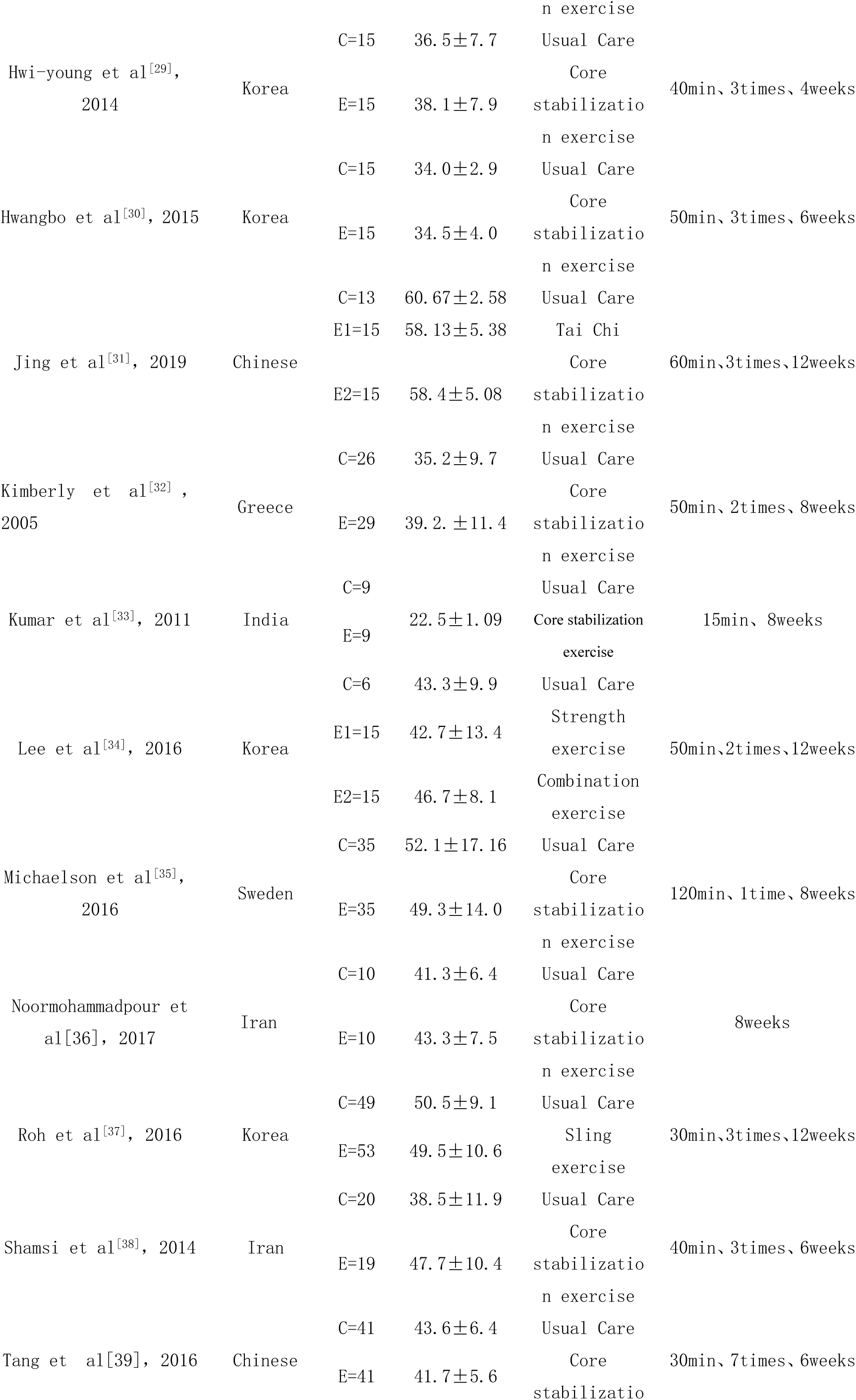
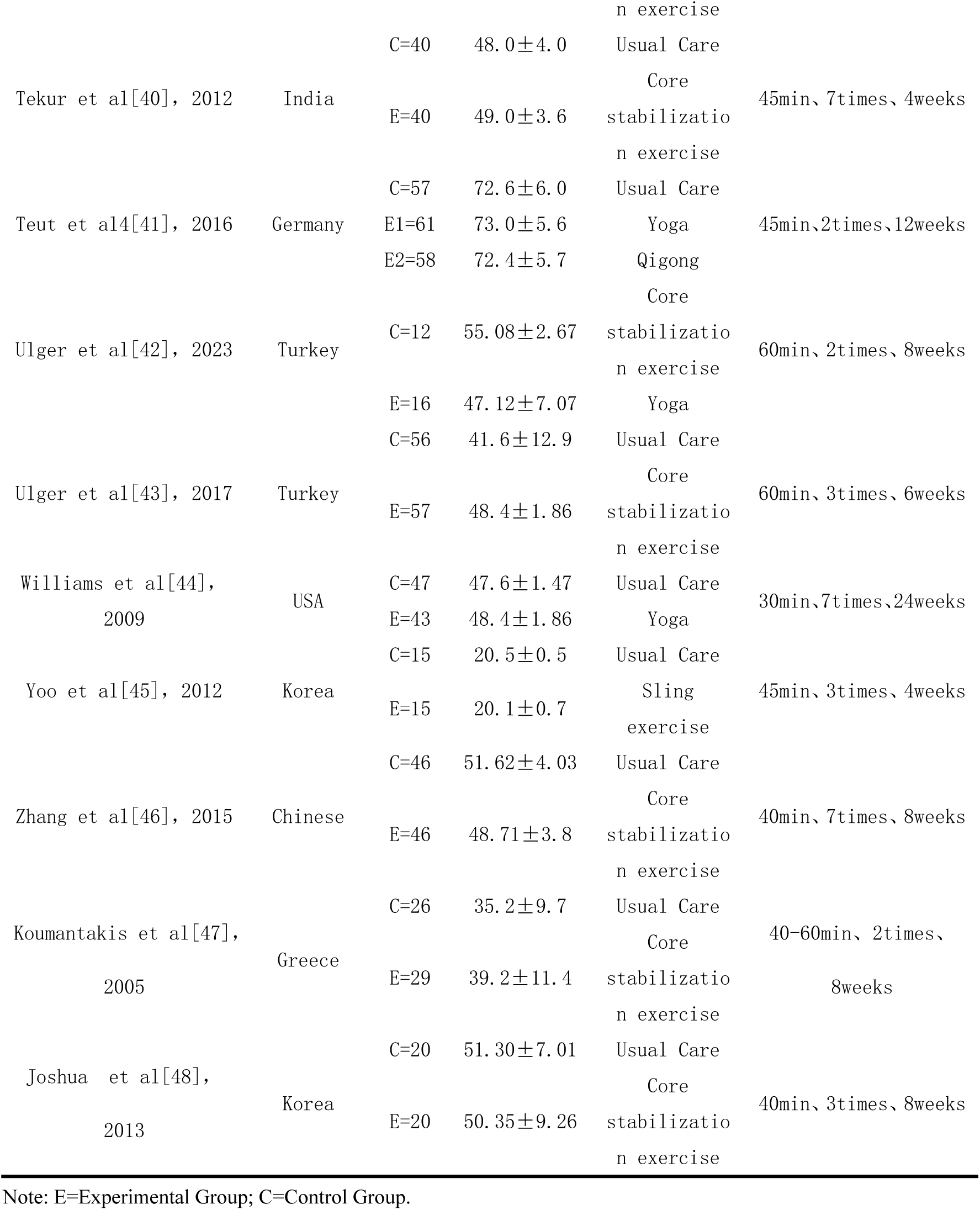
Basic characteristics of the included literature.

## 3. Network Meta-Analysis Results

### 3.1 Network Evidence Map

Figure 2 displays the network relationship diagram for 12 intervention measures affecting individuals with chronic nonspecific low back pain. Connections between the nodes indicate direct comparisons between two interventions, while the absence of a line indicates no direct comparison between them. The size of the circles represents the sample size included for each intervention method, and the width of the lines indicates the frequency of comparisons between the two interventions.

### 3.2 Results of the Network Meta-Analysis

The 26 studies reported on the efficacy of different exercise modalities for chronic nonspecific low back pain. Yoga [SMD = −1.71 (−2.93, −0.49), P < 0.05] and core stability training [SMD = −0.81 (−1.44, −0.18), P < 0.05] were significantly more effective than the control group. However, combined training [SMD = −1.48 (−4.03, 1.08), P > 0.05], Pilates [SMD = −0.55 (−3.00, 1.89), P > 0.05], Qigong [SMD = −1.23 (−3.88, 0.91), P > 0.05], suspension training [SMD = −1.40 (−2.82, 0.02), P > 0.05], Swiss ball exercises [SMD = −0.88 (−3.12, 1.36), P > 0.05], strength training [SMD = −1.36 (−3.91, 1.19), P > 0.05], perturbation therapy [SMD = −0.39 (−2.82, 2.04), P > 0.05], sit-up exercises [SMD = 0.09 (−2.82, 2.04), P > 0.05], and Tai Chi [SMD = −2.13 (−4.32, 0.06), P > 0.05] did not show statistically significant differences compared to the control group (see Table 2).

**Table 2.**
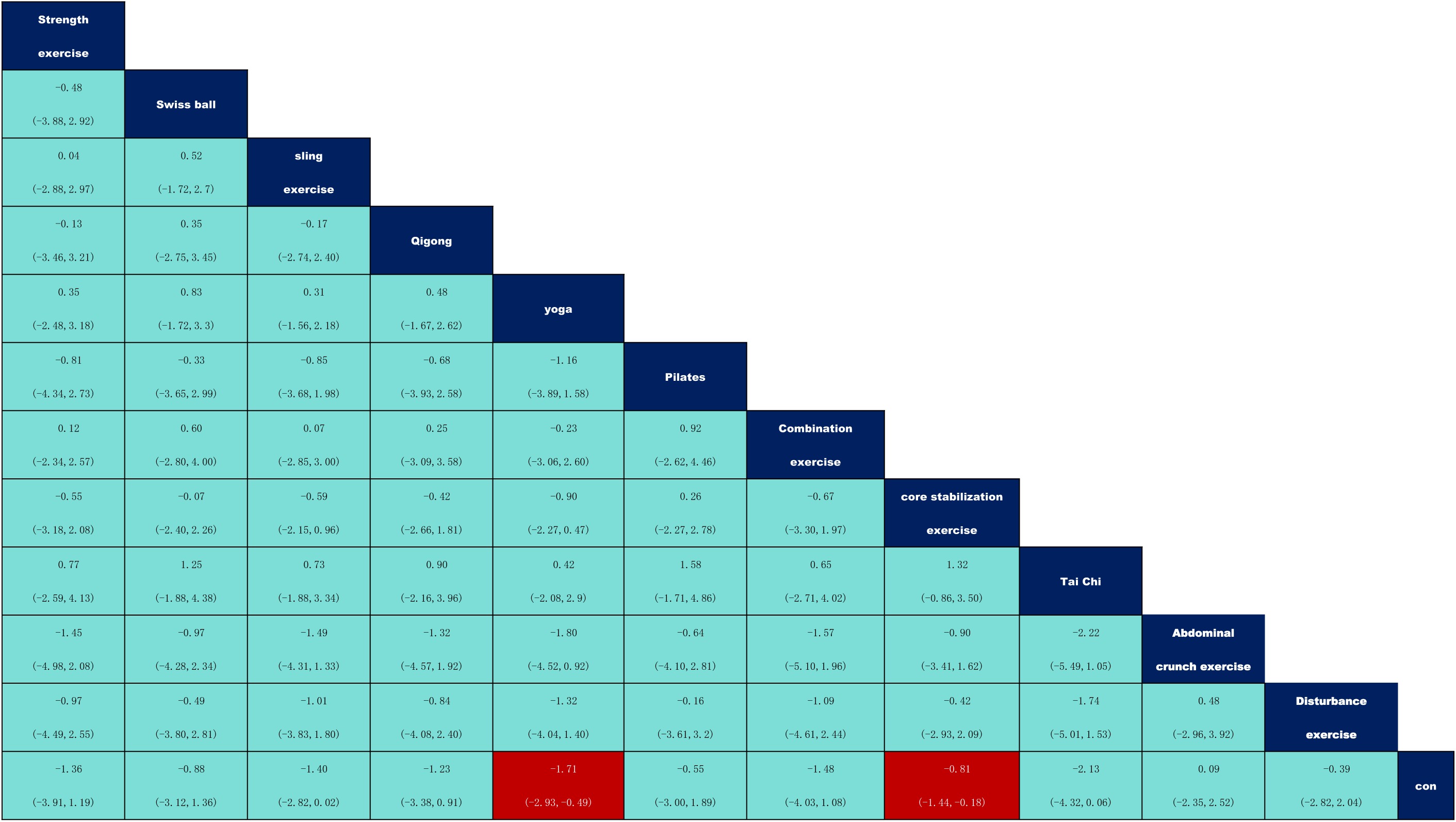
The results of cross comparison among interventi.

### 3.3 Ranking of Intervention Effectiveness

Based on the Surface Under the Cumulative Ranking (SUCRA) methodology, the potential ranking of effectiveness for interventions addressing chronic nonspecific low back pain among 12 different measures is as follows: Tai Chi (SUCRA = 77.4) > Yoga (SUCRA = 72.1) > Suspension Training (SUCRA = 63.0) > Combined Training (SUCRA = 61.6) > Strength Training (SUCRA = 59.2) > Qigong (SUCRA = 57.5) > Swiss Ball (SUCRA = 48.0) > Core Stability Training (SUCRA = 44.8) > Pilates (SUCRA = 40.5) > Perturbation Therapy (SUCRA = 34.5) > Sit-up Exercises (SUCRA = 23.4) (see Table 3).

**Table 3.** SUCRA values and effect sizes for effectiveness of each intervention.

| Intervention | SUCRA | Optimal<br>Probability (%) | Ranking |
| --- | --- | --- | --- |
| Taichi | 77.4 | 33.6 | 1 |
| Yoga | 72.1 | 8 | 2 |
| Sling exercise | 63 | 4.7 | 3 |
| Combination exercise | 61.6 | 13.8 | 4 |
| Strength exercise | 59.2 | 12.6 | 5 |
| Qigong | 57.5 | 11.2 | 6 |
| Swiss ball | 48 | 6.3 | 7 |
| Core stabilization exercise | 44.8 | 0 | 8 |
| Pilates | 40.5 | 4.9 | 9 |
| Disturbance exercise | 34.5 | 3 | 10 |
| Abdominal crunch exercise | 23.4 | 1.9 | 11 |
| Con | 18 | 0 | 12 |

### 3.4 Detection of Publication Bias

According to Figure 3, the included studies are mostly symmetrically distributed around the zero line, with the majority of points located in the upper part of the funnel, and only a few points falling outside the funnel plot. Overall, the results indicate a low possibility of publication bias in this study. The Egger’s method was used to test for publication bias, yielding a P-value of 0.161, suggesting no significant publication bias. However, the interpretation of these results should still be approached with caution.

## 4 Discussion

This study through network meta-analysis reveals that Tai Chi is more effective than other exercise interventions in alleviating chronic nonspecific low back pain in adults. This finding diverges from previous literature, which indicated that Pilates was the best exercise choice for improving chronic nonspecific low back pain in adults[49]. However, the SUCRA ranking results from this study place Tai Chi above combined exercises, Pilates, Qigong, suspension training, Swiss ball exercises, strength training, perturbation therapy, sit-up exercises, and yoga in terms of effectiveness. This suggests that Tai Chi could be considered a more effective strategy for treating and preventing chronic nonspecific low back pain. The unique effectiveness of Tai Chi in back pain intervention may be closely related to its emphasis on “slowness.” Practitioners gradually deepen their perception of subtle body changes through slow and coherent movements, enhancing proprioception. This heightened bodily awareness not only improves the coordination of muscle tension and relaxation but also focuses on joint stability and flexibility, as well as the overall flow and balance of qi and blood[50, 51].

Currently, there is no unified conclusion on the specific mechanisms by which physical activity improves and controls chronic nonspecific low back pain, but numerous studies consistently show that physical activity plays a significant role in reducing chronic nonspecific low back pain[52–55]; Semrau et al. observed significant pain relief in patients after an eight-week exercise intervention[56]; similarly, research by Zhu et al. further supports the positive impact of physical activity on reducing back pain[57]. The two major mechanisms universally accepted by the academic community are the neurophysiological mechanism and the vascular mechanism, which provide robust support for explaining the principles behind exercise interventions. (1) Neurophysiological mechanism: Numerous studies indicate that during exercise, the hypothalamus and pituitary gland of vertebrates release amino compounds and other neurotransmitters that have significant analgesic effects, reducing patients’ pain and alleviating discomfort caused by chronic nonspecific low back pain[58–60].Furthermore, exercise can regulate the activity of brain areas associated with pain perception, such as the parietal and frontal lobes, enhancing the brain’s capacity to process pain information and reducing the sensitivity of chronic low back pain patients to pain, thereby improving their quality of life[61, 62]. Exercise also significantly increases the secretion of neurotrophic factors, which play a crucial role in neuronal repair processes, helping to alleviate pain and promote the repair of damaged nerves[63]. (2) Vascular mechanism: Studies indicate that chronic nonspecific low back pain often accompanies local blood circulation obstruction and tissue hypoxia[64]. Prolonged sitting, standing, and improper exercise can hinder back muscle and blood circulation, exacerbating back pain [65].However, exercise can promote blood flow in back muscles and soft tissues, improving circulation. Specific exercise patterns can promote rhythmic muscle contractions, thus dilating blood vessels, improving blood supply and waste disposal, and alleviating muscle fatigue and pain[66–68].

In the realm of neurophysiology, Tai Chi emphasizes precise muscle control and the coordination of movements, which requires the brain to finely regulate the intensity and sequence of muscle contractions [62, 69]. This precise neural regulation enhances the nervous system’s response speed and sensitivity to movement, optimizes neural conduction pathways, and reduces errors in neural transmission [70].Additionally, Tai Chi enhances proprioceptive input to the lumbar and lower limb joints, restoring joint stability and effectively alleviating pain caused by joint instability[71, 72]. Chronic nonspecific low back pain is often associated with neural adhesions, which are typically due to chronic inflammation or poor posture limiting neural mobility and increasing pain signal transmission[73–75]. Tai Chi alleviates pain by increasing the range of neural mobility, reducing neural adhesions and compression [76]. Further research suggests that Tai Chi practice can regulate gene expression related to inflammation, reducing the activity of key inflammatory factors such as NF-κB, thus diminishing chronic inflammatory responses and associated pain [77, 78]. Moreover, Tai Chi enhances the secretion of endogenous analgesics like endorphins, providing emotional relief and a sense of pleasure from movement, thereby becoming an effective non-pharmacological treatment option[79].

The vascular mechanism further indicates that Tai Chi improves local blood circulation, accelerating the clearance of inflammatory products, reducing inflammatory responses, and alleviating pain[62, 80, 81]. More efficient blood circulation not only enhances the nutrient supply to muscles but also speeds up the elimination of metabolic waste, reducing pain caused by fatigue [82]. This improvement in blood circulation helps increase vascular elasticity, reducing the risk of arteriosclerosis and vascular stiffness[80]. Enhanced vascular elasticity can decrease the resistance in lumbar and lower limb vessels, increasing blood flow and thereby relieving chronic nonspecific low back pain[82].Additionally, emotional stress often exacerbates pain symptoms; negative emotions can lead to overactivation of the sympathetic nervous system, further causing muscle tension and poor blood circulation. Tai Chi, with its slow movements and deep breathing techniques, significantly reduces the activation level of the sympathetic nervous system, decreases the secretion of stress hormones like adrenaline, and helps patients achieve deep relaxation on both physiological and psychological levels[83].Finally, prolonged Tai Chi practice can enhance the muscular strength of cardiac tissues, increase the elasticity of the heart valves, improve cardiac output, return blood volume, and myocardial oxygen reserve capacity, significantly improving the cardiac blood supply system[84]. These changes help provide more blood and oxygen to the lumbar muscles and bones, promoting the repair and regeneration of lumbar tissues, thereby alleviating back pain.

## 5 Comprehensive Implications for Research and Clinical Practice Research Implications

### 5.1 Implications for research

Network meta-analysis enables indirect treatment comparisons (ITC) by quantifying and ranking different intervention measures. This approach allows us to clearly define the differences in effectiveness among various types of exercises in improving chronic low back pain in adults. By comparing the impact sizes of each type of exercise, we can identify the most effective exercise modality. This method facilitates the design of more precise randomized controlled trials for future research, ensuring that interventions are targeted and based on robust evidence. This strategy not only enhances the reliability of research findings but also contributes to the development of tailored treatment plans that are more likely to yield significant benefits for patients with chronic low back pain.

### 5.2 Implications for clinical practice

The results of this network meta-analysis provide a solid evidence base for guiding the selection of exercise interventions for adults with chronic low back pain. We found that Tai Chi is the most effective form of exercise for improving chronic low back pain in adults. We hope these findings will serve as valuable decision-making information for educators and experts who develop exercise guidelines. This should aid in formulating more targeted and effective strategies to manage and alleviate chronic low back pain through non-pharmacological interventions.

## 6. The limitations of this study

This study also has certain limitations: First, the study focuses only on exercise interventions and does not include other potential interventions (such as pharmacological treatments, psychological interventions, etc.), which may limit the applicability of the results. Lastly, because self-reported pain is highly subjective, these data may be affected by subjective perceptions changing over time, potentially impacting the accuracy of the data and thus the stability of the results. These limitations inevitably have a negative impact on the accuracy of the study conclusions, so the findings should be cautiously generalized.

## Data Availability

All relevant data are included in the manuscript and its Supporting Information files.

---

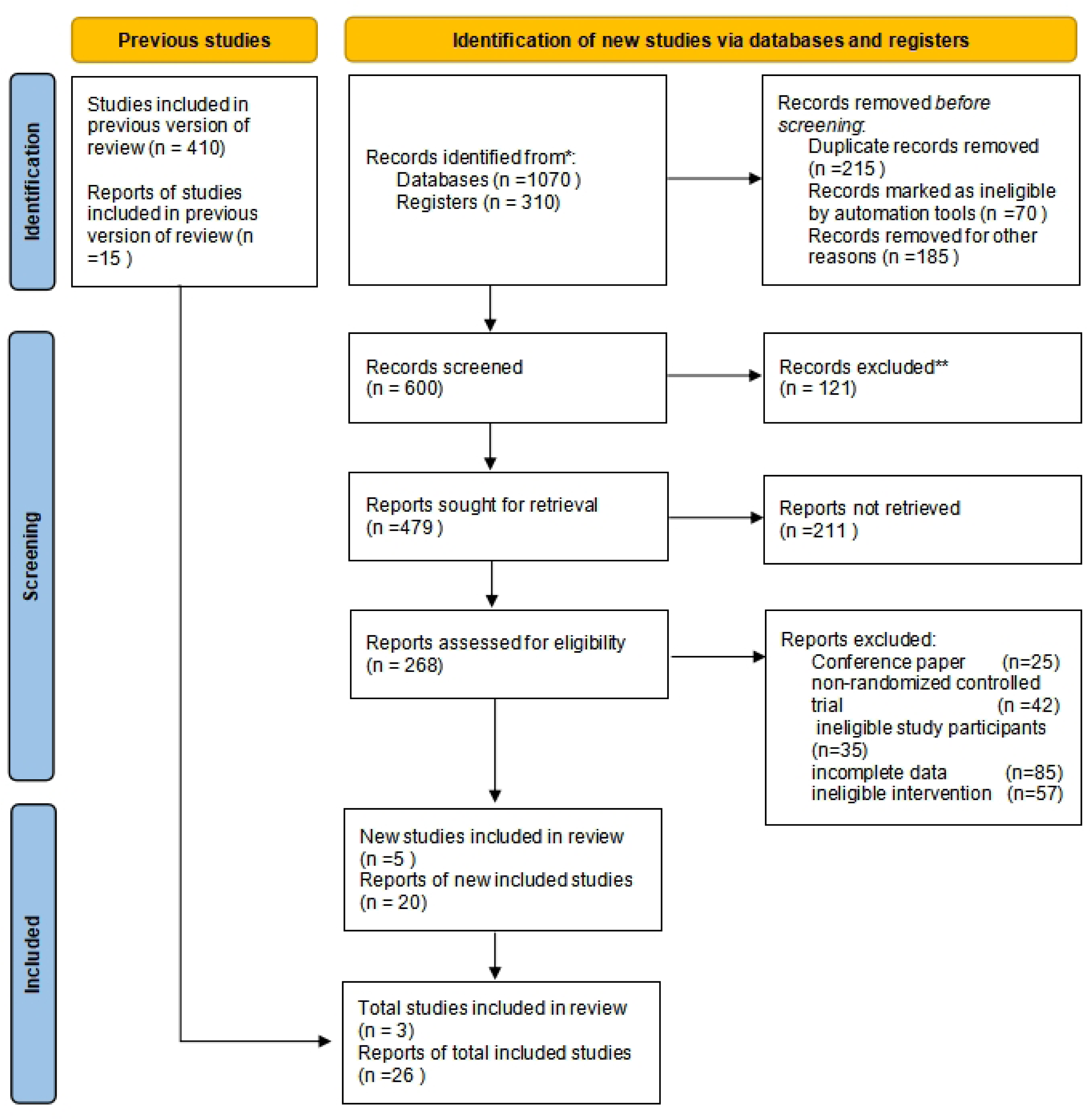

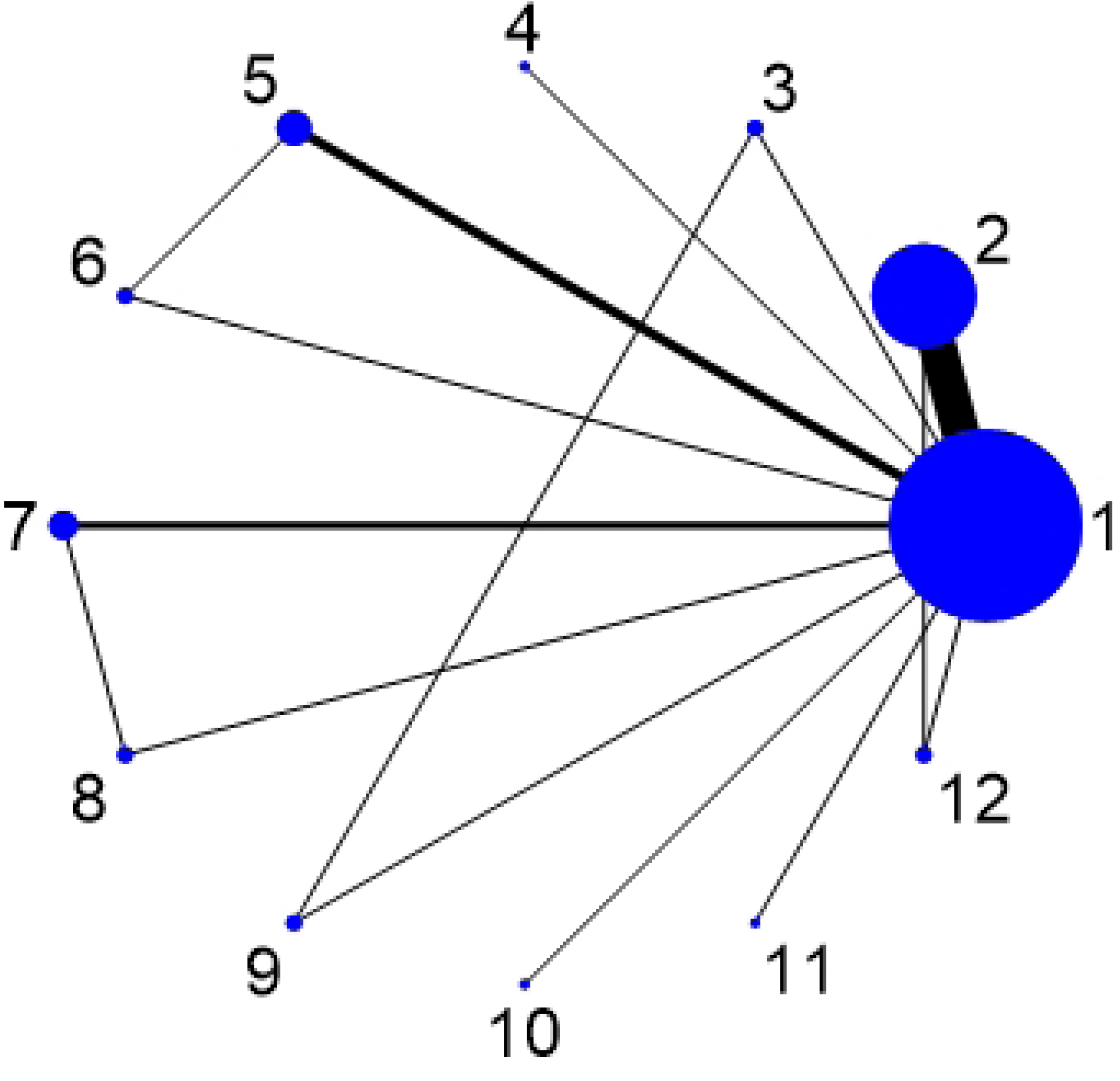

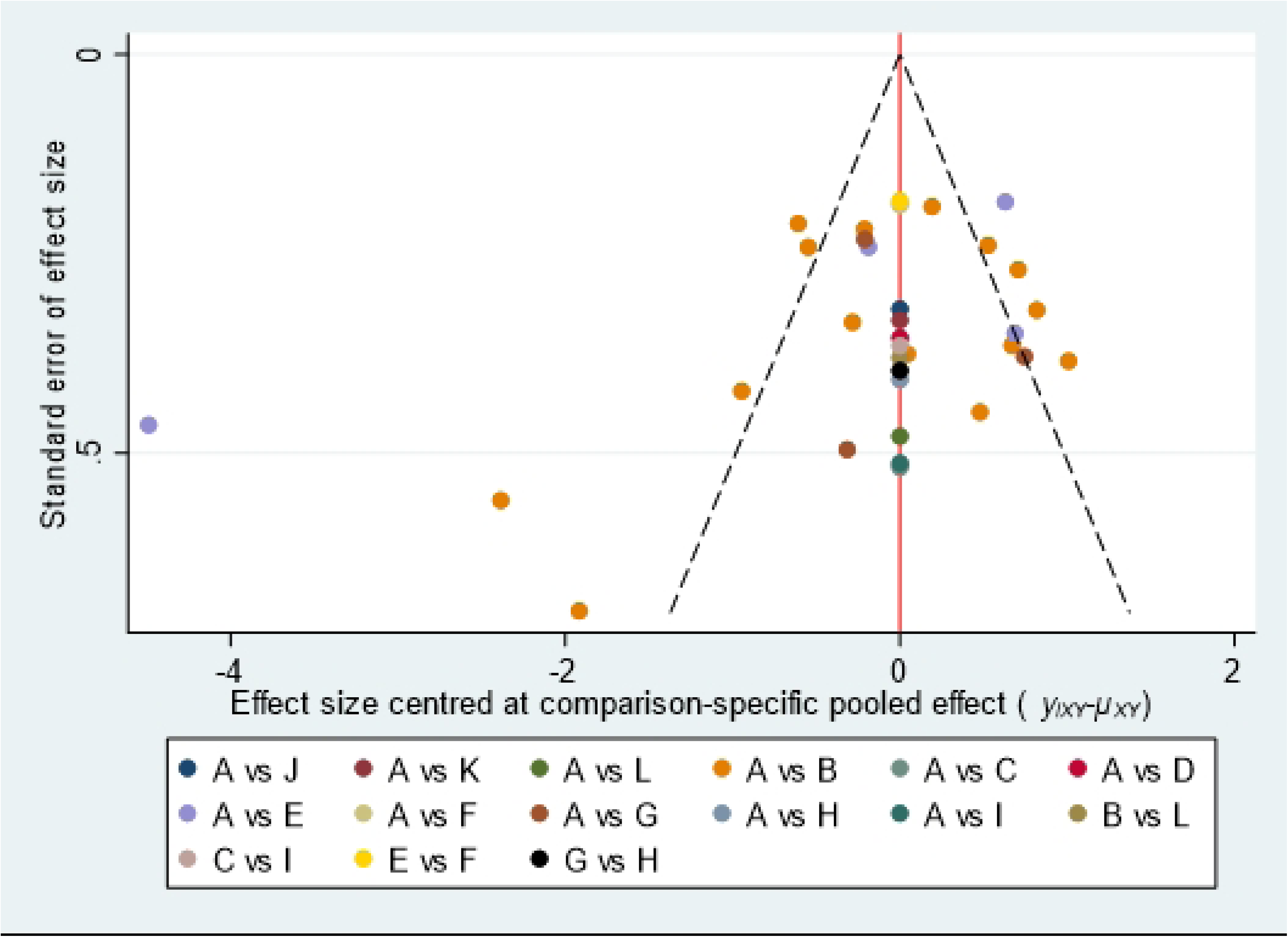

